# Understanding health innovation adoption: A realist evaluation of pulse oximeter implementation in primary care for children under five in four West African countries

**DOI:** 10.1101/2025.01.26.25321135

**Authors:** Sarah Louart, Habibata Balde, Abdourahmane Coulibaly, Abdoua Elhadji Dagobi, Kadidiatou Kadio, Gildas Boris Hedible, Valériane Leroy, Emilie Robert, Valéry Ridde, AIRE study group

## Abstract

**Introduction:** Hypoxemia is an important contributor to child mortality, particularly in low-resource settings where diagnostic tools are scarce. The AIRE project introduced pulse oximeters (POs) into 202 primary healthcare centres (PHCs) in Burkina Faso, Guinea, Mali, and Niger, integrating them into the Integrated Management of Childhood Illness (IMCI) guidelines. This initiative aimed to strengthen diagnostic capacities for identifying hypoxemia and to improve care management for critically ill children under five. This study examined how healthcare workers (HCWs) adopted POs and explored the contexts and mechanisms influencing their adoption.

**Methods:** We conducted a realist evaluation to analyse adoption patterns, focusing on interactions between the Intervention, Contexts, Actors, Mechanisms, and Outcomes (ICAMO configurations). Data collection included 299 interviews with HCWs, patients’ families, and institutional actors, conducted in 16 selected PHCs, at the institutional level, and in district hospitals, complemented by site observations. Analysis was performed using NVivo software, identifying ICAMO configurations as demi-regularities to explain variations in PO use and adoption.

**Results:** Training enabled HCWs to recognize the utility of POs, further motivating their use. Support-focused supervision fostering a sense of support, while control-focused approaches sometimes resulted in mechanical use driven by external pressure. In contexts of high workloads and children’s agitation, difficulties in using POs were observed. In settings with limited diagnostic tools, POs increased HCWs’ diagnostic confidence, encouraging adoption and improving decision-making. Observing or knowing the benefits of POs on children’s health provided HCWs with a sense of relief and pride, further reinforcing PO adoption. However, structural barriers and challenges related to institutional adoption may threaten long-term use.

**Conclusion:** This study highlights the necessary conditions for adopting POs in PHCs. While widely used by healthcare workers, addressing challenges related to training, supply chain logistics, and referral systems to hospitals is essential to ensure long-term sustainability and improve child health outcomes.

**What is already known on this topic:**

- Severe hypoxemia significantly contributes to child mortality, especially in low-resource settings lacking adequate diagnostic tools.
- Pulse oximeters (POs) are effective in detecting hypoxemia and guiding referrals, particularly when integrated into the Integrated Management of Childhood Illness (IMCI) guidelines.

**What this study adds:**

- Using a realist evaluation approach, this study identified diverse responses to PO implementation, explaining the reasons for non-use, mechanical use, use, and adoption. POs appear to be predominantly adopted at an individual level.
- Findings highlight the critical role of boosting confidence in health care providers’ diagnostics, as well as the sense of relief and pride derived from participating in the care of children, to support adoption.
- Structural and institutional challenges, including training, supply chain issues, device maintenance, and referral system inadequacies, were found to potentially hinder the sustainability of PO adoption.

**How this study might affect research, practice, or policy:**

- The study provides valuable insights into the contexts and mechanisms that facilitate or hinder the adoption of POs, which may also inform the integration of other diagnostic tools into primary healthcare systems.
- Policymakers and healthcare implementers can utilize these findings to design tailored strategies to implement similar interventions in other contexts.

## Introduction

Infectious diseases, particularly respiratory infections are leading causes of child mortality [1,2], especially in sub-Saharan Africa, where socio-economic factors such as poverty and inadequate home ventilation contribute to their high prevalence [3]. West Africa has the highest child mortality rates under five, largely due to infectious diseases. However, efforts and resources to address this public health issue remain insufficient [4].

Severe hypoxemia, defined as a blood oxygen concentration below 90%, is a key indicator of the severity of acute lower respiratory tract infections [5] but also a marker of severity for other common conditions like malaria and malnutrition [6]. In low- and middle-income countries, hospital studies report significant prevalence rates of hypoxemia, varying according to age, disease type, and illness severity [7–9]. This severe hypoxemia substantially increases the risk of death in children [5]. However, clinical symptoms alone are insufficient to reliably detect hypoxemia, underscoring the interest of devices that measure oxygen saturation for identifying children requiring urgent care [10,11], including oxygen therapy [12,13]. International calls have long advocated for improved access to oxygen and diagnostic tools like pulse oximeters (PO) [13], which measure oxygen saturation and help detect hypoxemia cases.

To enhance child healthcare, the World Health Organization (WHO) introduced the Integrated Management of Childhood Illness (IMCI) strategy in the 1990s, aiming to prevent and treat severe childhood illnesses in low-resource settings [14]. In 2014, the IMCI clinical guidelines were updated to include specific recommendations for respiratory diseases, advocating for the use of PO “when available” and the transfer to facilities equipped to administer oxygen when oxygen saturation falls below 90% [15]. However, access to PO and oxygen in low-income countries remains inadequate [16], especially in primary care settings [17], limiting the potential impact of this recommendation.

Although studies have demonstrated the effectiveness of PO for identifying critical cases that could go unnoticed with IMCI alone [18,19], evidence on its impact at the decentralized level remains scarce. A study in Uganda is one of the few to examine this aspect [20]. Furthermore, there is a significant gap in qualitative research exploring the processes of PO implementation and adoption, particularly in decentralized primary care settings [21]. Recently, a realist evaluation in Nigeria provided valuable insights, revealing that PO adoption in these settings was both limited and uneven [22].

To tackle these challenges, the AIRE project was implemented in four West African countries: Burkina Faso, Guinea, Mali, and Niger. Led by a consortium of NGOs, this project aimed to strengthen diagnostic capacities for severe hypoxemic cases at the primary care level by introducing the routine use of PO during IMCI consultations. A multi-faceted study was conducted to understand the introduction of PO in these settings [23]. A key element in disseminating innovations is understanding the factors influencing their adoption (or non-adoption) [24]. Given the complexity of implementing innovations in primary health care centres (PHCs), a realist approach [25] was used to explore the contexts and mechanisms that facilitate or hinder the adoption of PO by healthcare workers (HCWs). This study sought to answer the overarching question: how does the AIRE project work (or not), for whom, in which specific contexts, and why? It further explored how HCWs responded to the intervention, which contextual elements shaped these responses, and what effects these responses produced on the use and adoption of PO.

## Methods

### Intervention and research sites

The AIRE intervention was implemented from July 2019 to December 2022 in two health districts within each of the four countries: Burkina Faso, Guinea, Mali, and Niger (Figure 1), encompassing 202 PHCs and 8 district hospitals. The IMCI guidelines were primarily utilized in paper format, except in Burkina Faso and one district in Mali, where electronic IMCI was employed as part of the Integrated e-Diagnostic Approach (IeDA) program [26]. Project activities included training in PO use and refreshing HCWs in IMCI, distributing POs, conducting supervision visits, and strengthening care capacities at PHCs (basic medicines and medical equipment) and district hospitals (POs and oxygen concentrators).

**Figure 1.**
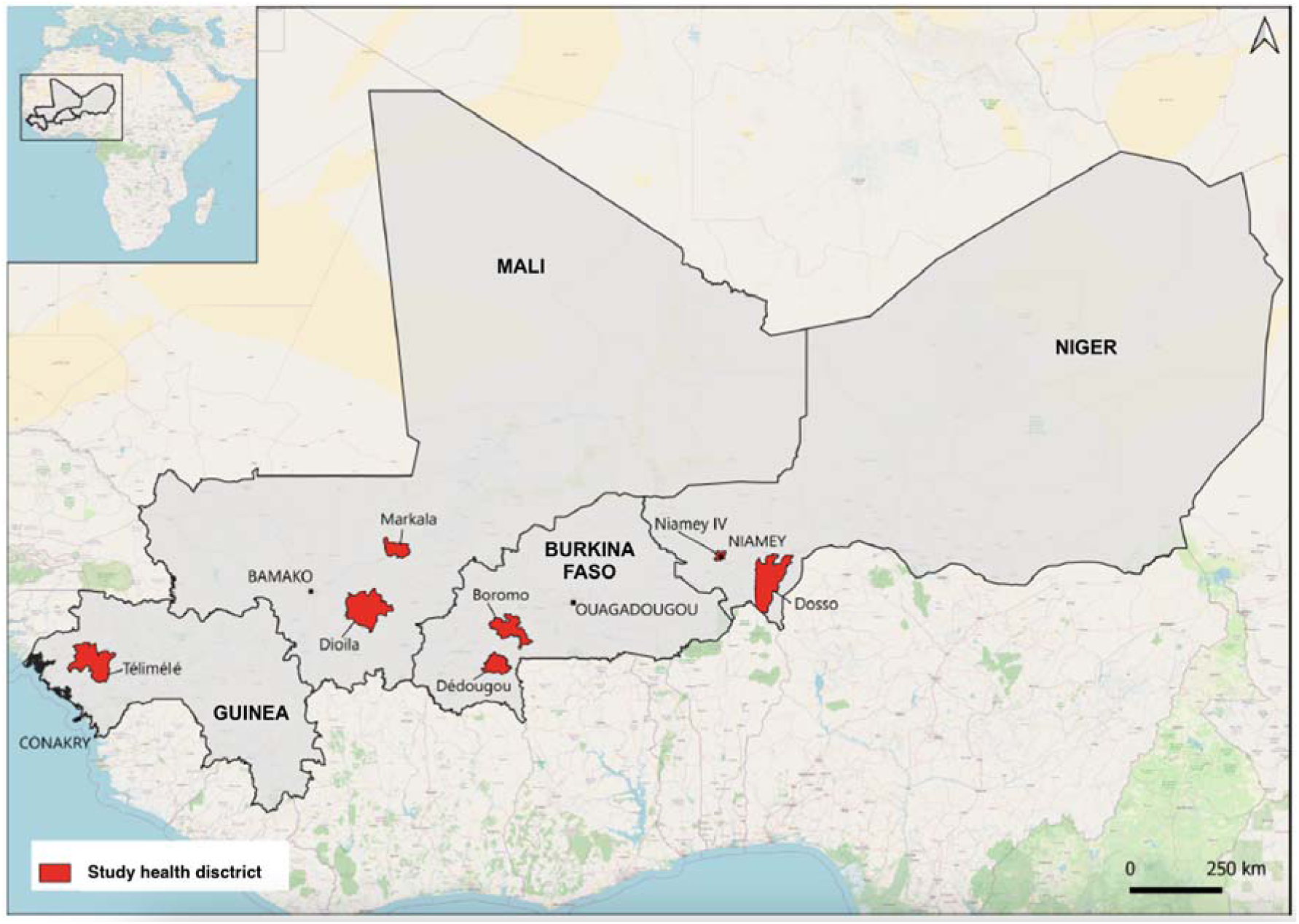
Countries and health districts involved in the AIRE study

The research was conducted primarily in 16 selected PHCs (four per country), designated as sites 1 through 4 in each country. Additional investigations occurred at hospital and institutional levels, including district teams and ministries. Details on the intervention, research site selection criteria, and eligibility criteria for children’s PO use are outlined in the study protocol [23]. Characteristics of the sites are described elsewhere [27].

### Study design

To gain an in-depth understanding of HCWs’ adoption or non-adoption of PO, we conducted a realist evaluation [25]. This approach seeks to understand how and why an intervention produces specific effects by exploring underlying mechanisms— namely, the reasoning and reactions of individuals in response to resources provided through the intervention [28]. These mechanisms are triggered only in specific contexts, requiring an analysis of the dynamic interactions between context, mechanisms, and observed outcomes. To model this dynamic, realist evaluation employs Context-Mechanism-Outcome (CMO) configurations as a heuristic tool, facilitating the development of analytical and robust explanations of results. We began by formulating our hypotheses using Intervention-Context-Actor-Mechanism-Outcome (ICAMO) configurations to specify the intervention and target actors of our study (PHC HCWs) [29]. As we moved toward a higher level of abstraction, we employed CMO configurations to develop the middle-range theory (MRT).

The realist evaluation process begins with reconstructing the program theory, defined as the underlying assumptions about how an intervention is intended to work and the impacts it is expected to have [30]. These initial assumptions are gradually refined, aiming to develop a middle-range theory (MRT) [31]. Realist evaluation thus favours theoretical generalization [32], supporting the formulation of practical and contextualized recommendations for policymakers by progressively refining the program’s initial theory.

Our realist evaluation design is illustrated in Figure 2. We organized three research workshops to guide the evaluation: the first framed the research and refined the initial intervention model through discussions with coordination teams at both country and international levels, while the second and third focused on adjusting and refining hypotheses, leading to the formulation of 26 theoretical hypotheses. These hypotheses were developed based on preliminary field data, scientific literature— including realist studies on PO use in Nigerian hospitals [33], a realist review on access to oxygen, and extensive literature on innovation adoption [34–36]. The process of refining the program theory is detailed in Appendix 1.

**Figure 2.**
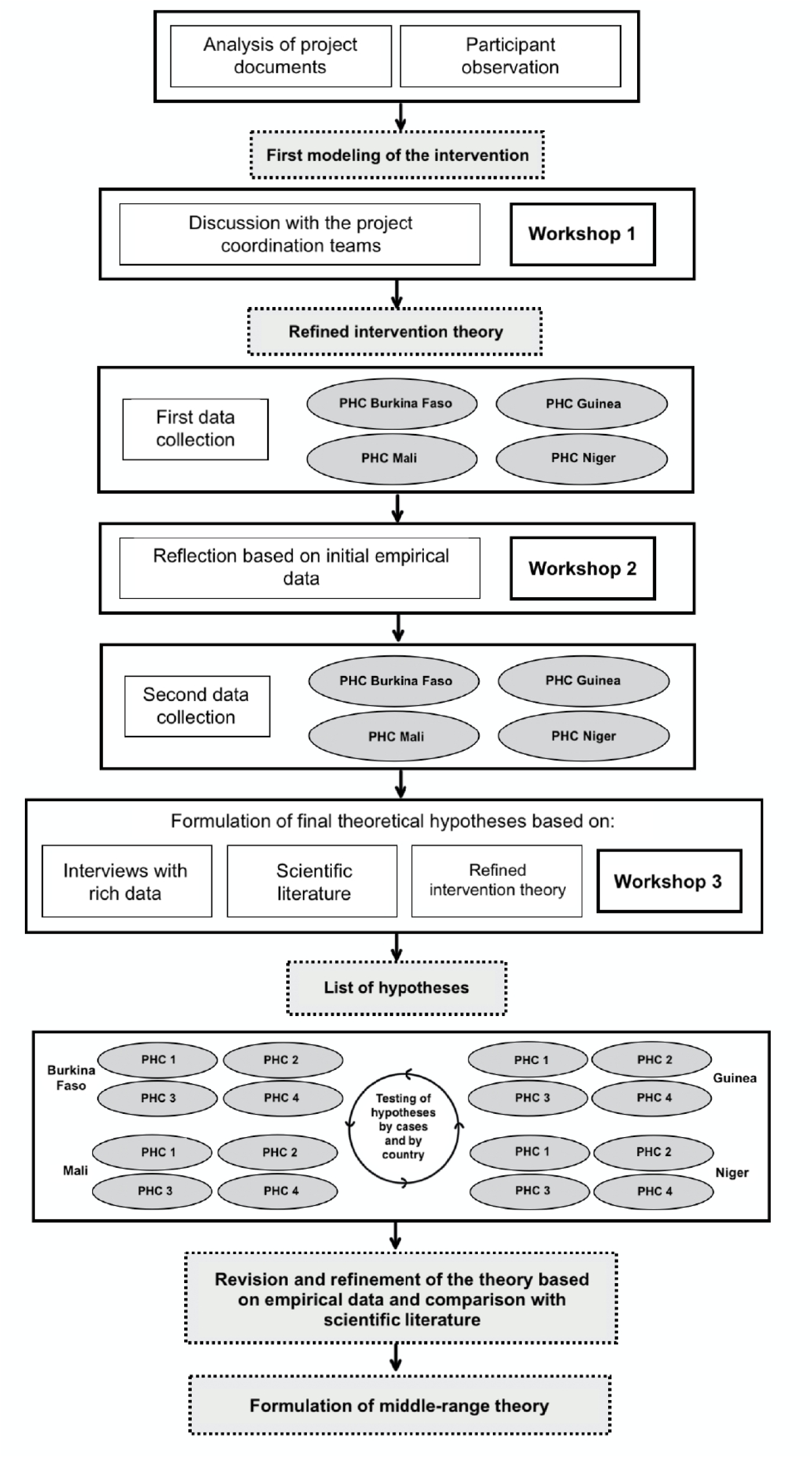
Realist evaluation design of the AIRE Project

This article follows the RAMESES guidelines for reporting realist evaluations to ensure transparency and rigor [37].

### Data collection

We conducted two rounds of data collection (Figure 3). The first (T1) occurred in November and December 2021, approximately six months after introducing PO in the PHCs. The second (T2) took place in October and November 2022, at the end of the project’s main activities. Experienced national qualitative researchers conducted in-depth individual interviews after obtaining written consent. Interviews were conducted in French or local languages, depending on the location and participants.

**Figure 3.**
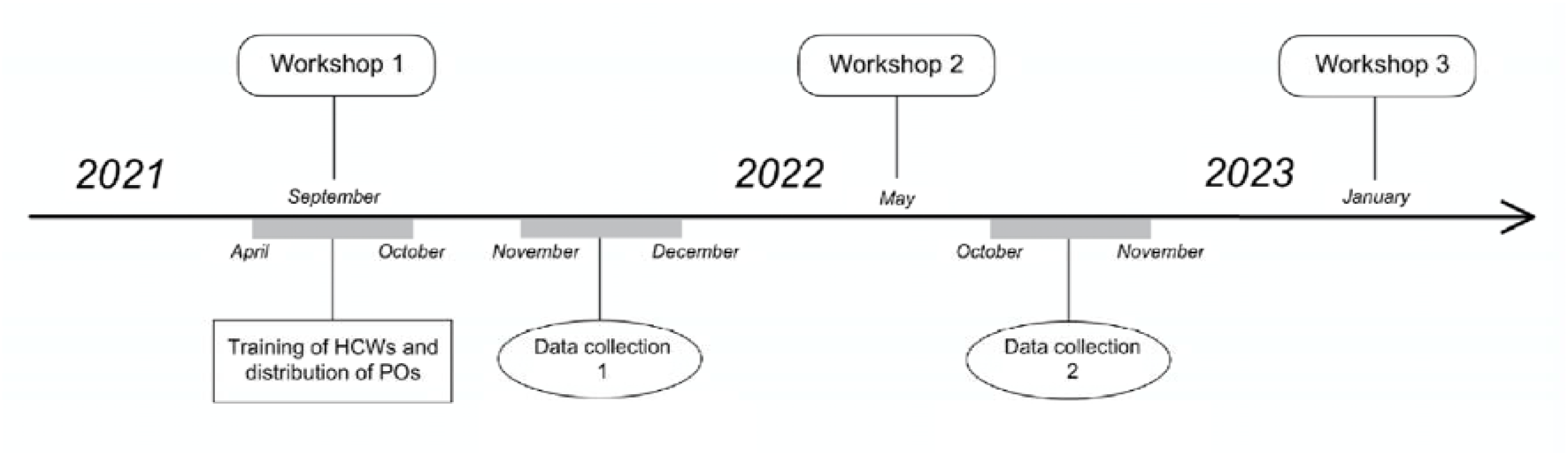
Timeline of the study

In both phases, we employed exhaustive sampling for HCWs consulting children under five at PHCs and available during the survey. For other groups, including families of the children, HCWs at district hospitals, and individuals involved in project monitoring at the institutional level, we used purposive sampling [23]. Additionally, observations were conducted in PHCs, including consultation observations.

### Data Analysis

The interviews conducted with HCWs in PHCs serve as the primary data source for this study. Additional interviews with other participants and observations carried out in PHCs, were used for data triangulation, further strengthening the realist analysis.

Our analysis focused on identifying four main effects of the intervention: use, non-use, unsustainable use (mechanical, circumstantial, or temporary), and adoption of the PO. Use encompasses any application of the PO, whether systematic, anecdotal, mechanical, or performed under constraint. It includes both instances that may progress toward adoption and those that remain unsustainable. Importantly, use is considered a necessary step toward adoption, as specific configurations associated with use can generate feedback loops that ultimately support adoption. Adoption implies a voluntary and sustained integration of the PO into HCWs’ practices.

Data analysis followed an iterative process guided by realist methodology. Hypotheses were examined across all interviews and progressively refined based on emerging findings. To ensure rigor and reliability, double coding was conducted using Nvivo12. An approach by case studies was adopted, with cases selected for their distinct characteristics to guide data collection. Comparisons were then conducted within and across cases to identify patterns, refine interpretations, and strengthen the analysis [32]. The list of hypotheses was further revised and refined by integrating empirical findings with existing literature. This iterative process ultimately led to the formulation of a MRT.

### Ethical Approvals

As detailed in the study protocol, this research received approval from the ethics committees of the four participating countries, as well as from the ethics committees of WHO and Inserm [22].

### Patient and public involvement

Data collection was authorized by ethics committees and ministries of health. The study participants were not directly involved in the analysis, interpretation of results, or manuscript preparation.

## Results

During the two rounds of data collection, 299 interviews were conducted with various target populations. Table 1 presents the distribution by timeframe and country. Table 2 presents the different configurations of our MRT, developed based on the demi-regularities identified in the data, supported by examples of citations.

**Table 1.** Distribution of Interviews by data collection period, respondent groups, and country (N=299).

| Data collection period | Respondents | Countries |  |  |  | Total |
| --- | --- | --- | --- | --- | --- | --- |
|  |  | Burkina Faso | Guinea | Mali | Niger |  |
| T1 | HCWs at PHCs | 11 | 11 | 12 | 13 | **47** |
|  | Other PHC staff | 3 | 8 | 12 | 11 | **34** |
|  | Community health workers | 7 | 8 | 8 | 8 | **31** |
|  | Families of children | 16 | 16 | 16 | 11 | **59** |
| **Total T1 = 171 interviews** |  |  |  |  |  |  |
| T2 | HCWs at PHCs | 13 | 11 | 12 | 17 | **53** |
|  | Hospital health workers | 20 | 5 | 17 | 9 | **51** |
|  | Institutional level | 3 | 6 | 6 | 9 | **24** |
| **Total T2 = 128 interviews** |  |  |  |  |  |  |
| **Overall total = 299 interviews conducted** |  |  |  |  |  |  |

**Table 2.** Configurations of the Middle-Range Theory.

|  |  |
| --- | --- |
| **When the PO is introduced at the decentralized level, where technology is scarce, and in selected centers – as the project was implemented in specific districts rather than nationwide, with only some centers within these districts selected for the research (C), HCWs experience a sense of pride (M), a feeling of luck (M), an enhanced sense of legitimacy (M), and a pressure to meet expectations (M), which encourages them to use it (O).** | “When I use it, I feel like I’ve become more legitimate, like I’ve become a doctor, because before, it was the doctor’s job. You only see it in intensive care units. (...) Now, we feel proud. (...) The introduction of the PO has turned us into doctors” (Niger, T1, PHC 4)<br>“Since we’re a study site, that already makes a difference from other health sites (...) we’re held to good practices to make sure it goes well. (...) Being told that our site would be a study site was stressful. (...) For the health center, it’s a privilege for us to be part of the study. (...) If the research is successful, it’s a national pride, not just regional for the district.” (Burkina Faso, T1, PHC 2)<br>“Q: How do you feel each time you use the PO in your consultations? R: We’re proud. Q: Why? A: Because everyone who sees you using this machine trusts you.” (Guinea, T2, PHC 3) |
| **When HCWs are trained in the use of PO (C), they become aware of its utility and potential benefits (M),** | “Since the training, we have seen that it’s very important, because before we didn’t have a device that could really tell us if a child was in respiratory distress or not. We were treating them, I could say, ‘blindly’. But after the training, we |
| **which motivates them to use it (O).** | saw the importance of the PO, really, it's something very good." (Guinea, T1, PHC 1) |
| **When regular oversight is imposed on HCWs (C), they feel obligated to use the technology to meet expectations and avoid reprimands (M). This compelled use is often mechanical and driven by external pressure rather than a genuine belief in the tool's benefits, which may jeopardize sustainable use of PO (O). This obligation may also arise in contexts where usage is mandated by the digitization of IMCI.** | "At the very beginning, people weren't interested in the tool. But now, such cases are rare. At first, when you examine a child and refer them to the AIRE investigator, the investigator will ask the mother if the healthcare worker used something on the child's finger. If the mother says no, the investigator will personally get up, go with the mother, and confront you, saying the mother mentioned you didn't use the PO. I know this happened once or twice with a healthcare worker at the PHC here. Since that day, no mother comes with her child without the PO being used." (Burkina Faso, T2, PHC 1)<br>"The head of center emphasized that if someone fails to use the tool and misses a diagnosis, they will be held accountable. If you don't use it and a child dies under your care, you will be held responsible. She made this point very clear during our meetings and reiterated it at every single one." (Burkina Faso, T2, PHC 1)<br>"During supervisions, they check the registers and all the records... When we measure saturation, we're required to document it, and as soon as a supervision visit takes place, it's obvious they'll review the register to see if the PO is being used." (Niger, T2, PHC 1)<br>"Since we're using the IMCI-REC consultation, you can't even skip it. Because if you don't use the PO when the child is eligible... When you need to take the saturation, if you click 'no,' the REC will ask why: is it because the child isn't calm? Because the PO is malfunctioning? And so on. So, you need to answer all these questions before moving on. (...) With the REC, you can't move forward without taking the measurement first." (Burkina Faso, T1, PHC 1) |
| **When HCWs are actively and positively supported in their use of the PO, through regular guidance or encouragement that highlights its value (C), they feel supported (M) and recognize its relevance and usefulness (M). This motivates them to integrate the PO into their activities, thereby promoting a more voluntary use of the tool (O).** | "Supervision comes to galvanize us. Supervision is like moral support, an accompaniment. It encourages us to use the PO." (Burkina Faso, T2, PHC 1)<br>"That's what tells us the device is truly useful. They encourage us, they come to check, they supervise us." (Niger, T2, PHC 4). |
| **In contexts where patient management is complicated, and the workload is perceived as high (C), HCWs** | "It's impossible to use it on certain children because once the child starts crying, all the numbers the PO gives will be inaccurate. (...) There are some children who categorically refuse to have the cable attached to their toe or finger. (...) If I |
| **perceive the use of PO as an additional burden (M), leading to reluctance to use it optimally or, in some cases, rendering its use impossible (O).** | encounter this type of child every time during consultation, it can lead to discouragement.” (Mali, T1, PHC 1)<br>“Care slows down significantly, especially when we’re tired, and particularly here at [PHC 1], where patients only come at night. You’re exhausted, you’ve been working since morning until 6 or 7 p.m. (...) You’re tired, you want to prepare yourself, pray, wash, and they bring you children. And you absolutely have to use the pulse oximeter... Honestly, you can see there’s a bit of an overload... Yeah, it’s overwhelming.” (Niger, T1, PHC 1) |
| **In contexts where HCWs face challenges in making diagnoses due to material limitations (C), the use of PO enhances their confidence in themselves and their ability to make accurate diagnoses (M), encouraging its adoption (O).** | “Previously, we struggled with children here, doing things blindly because we had no way of truly knowing what their problem was. (...) I use the PO to feel good, to be confident in the outcome of the consultation. I do it to spare myself any doubt about the oxygen level in a child’s blood. The PO gives me something concrete and allows me to feel at ease and confident.” (Mali, T2, PHC 4) |
| **When HCWs are informed about the benefits of this technology or have observed its positive impact on patients’ health directly (C), it provides them with a sense of relief and pride (M), which further supports PO adoption (O).** | “The pulse oximeter has helped us progress a lot because we’ve truly seen its usefulness. (...) When we say that this device shows us that oxygen isn’t circulating in the blood, meaning the child needs to be referred, the mothers take them [to the hospital]. Their children are treated, and when they recover, they come back to thank us. They say they are very happy that their children have recovered.” (Niger, T2, PHC 1) |

We present the main configurations identified in the data, outlining the key contexts, mechanisms, and outcomes that explain the use and adoption of pulse oximeters in primary healthcare settings. These results are summarized in Figure 4.

**Figure 4.**
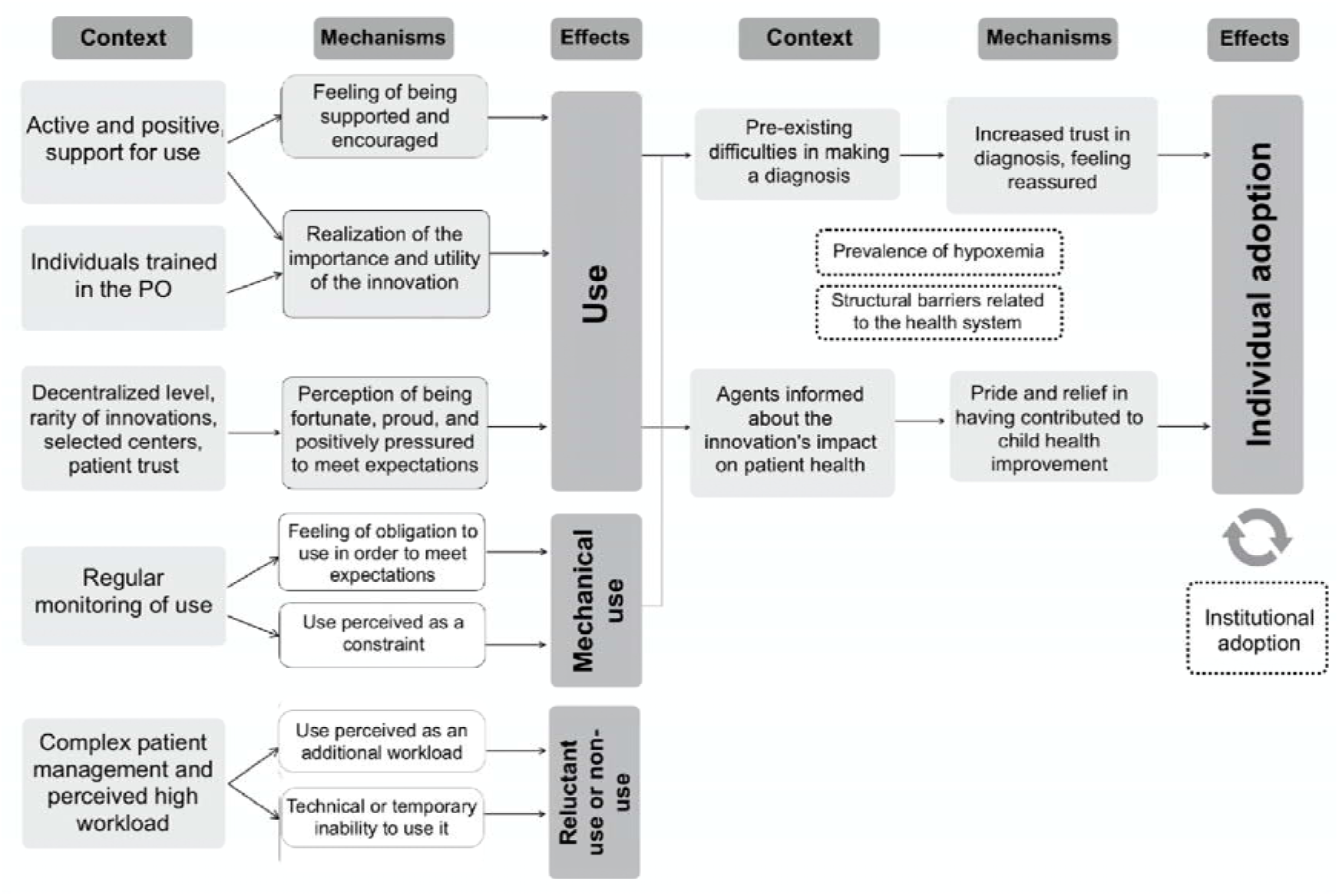
Middle-range theory of PO adoption

### PO use

The introduction of the PO at the decentralized level triggered a range of responses and mechanisms among PHC HCWs. First, in these settings, diagnostic technologies are typically reserved for higher levels of care and high-status professionals, such as doctors. Introducing the PO at the primary care level was thus perceived as a source of pride, enhancing HCWs’ sense of legitimacy in providing care. This perception was reinforced, particularly in Burkina Faso and Mali, by the sense of privilege associated with having the PO in selected centres, especially since other PHCs in the country did not receive the device.

Additionally, being part of a pilot project added pressure and motivation for HCWs to use the PO, as its potential scaling partly depended on the data collected in these research PHCs. Participation in the project fostered both a sense of pride and a responsibility to meet expectations and deliver positive outcomes. Some HCWs emphasized the need “to set an example” (Burkina Faso, T2, PHC2) or “not to disappoint” (Niger, T1, PHC3) project members.

This feeling of pride was also reinforced by HCWs’ perception of increased family trust and higher attendance at PHCs. In areas where PHCs often lack diagnostic tools, the PO enhanced the centre’s image, attracting more patients and strengthening trust as families appreciated the introduction of this new technology. This perception was further confirmed by interviews with families, who expressed greater confidence in the care provided. As HCWs noted, “the quality of the consultation is there, and it attracts patients” (Guinea, T1, PHC3) and “the PO is a tool that has created more trust between us and the patients” (Mali, T1, PHC1).

Another configuration underscores the key role of training in promoting PO use by raising awareness of its practical benefits. This effect became particularly evident during the second round of data collection, as the progressive training sessions conducted through the AIRE project enabled an increasing number of HCWs to recognize the value of the PO in their practice. Training sessions not only enhanced HCWs’ understanding of the PO’s value in refining diagnoses and detecting emergency situations related to hypoxemia, but also emphasized its role in facilitating referrals to hospitals where children could access oxygen therapy.

External motivation also influenced PO use, with two configurations emerging: control-focused supervision and support-focused supervision. Monitoring took various forms, including supervision by project and district teams, the presence of clinical investigators in PHCs, and oversight by PHC managers.

Support-focused supervision prioritized encouragement and guidance, fostering a more voluntary use of the PO. This approach helped HCWs feel supported and reinforced their understanding of the project’s importance. This support came from various sources. In some cases, the presence of clinical investigators was positively received. When supervision by the project was inclusive – involving all staff members rather than focusing solely on PHC managers – it also further promoted PO use.

Positive support from PHC managers was also highlighted by several HCWs. Managers who actively used the PO themselves were perceived as setting an example and “showing its importance to others” (Burkina Faso, T1, PHC3). This visible leadership encouraged other HCWs to view the PO as an essential tool, reinforcing their commitment to use it.

### Po mechanical use

In contrast, control-focused supervision emphasized performance monitoring and compliance. Clinical investigators, employed by the project and present daily in the PHCs for over a year, exerted a notable influence on PO use, particularly highlighted in Burkina Faso and Guinea. Investigators, tasked with collecting clinical data, actively pressured HCWs to use the PO by checking registers, insisting on its use, and reminding those who did not comply. As one HCW noted: “they put a lot of pressure on us” (Guinea, T1, PHC2).

In some cases, PHC managers also contributed to integrating PO into daily practices by exerting pressure to ensure its use. While this was not observed in Guinea, it was reported in nearly all other cases. In Niger, one HCW explained: “I assure you that she [PHC manager] monitors strictly! So, no one wants to be the source of any issues!” (Niger, T2, PHC3). This pressure often took the form of record checks, consultation monitoring, or reminders of responsibility.

Supervision was also sometimes perceived as a form of control, particularly during the second phase of data collection and in Burkina Faso. However, the perception of supervision varied depending on how it was implemented. In some cases, supervisors focused more on outcomes than support, leading HCWs to prepare to avoid exposing errors.

Additionally, the digitization of IMCI protocols sometimes compelled PO use. The electronic consultation register required oxygen saturation measurements to proceed with diagnoses, reinforcing PO use in PHCs implementing electronic IMCI.

### Challenges in using the PO

We also identified contexts that may hinder PO use. Children’s agitation emerged as the primary reason for non-use, particularly during the first phase of data collection. This agitation not only prolonged the time required to use the device but, in some cases, made its use entirely impossible. Observations conducted in PHCs confirmed these challenges, as HCWs were sometimes seen struggling to obtain readings on agitated children. Often linked to a general fear of healthcare facilities or the use of the malaria Rapid Diagnostic Test (mRDT) requiring a puncture, this issue prompted HCWs to adapt their practices. For example, some HCWs used the PO at the beginning of the consultation (before the mRDT) and employed various strategies to calm children and demonstrate that the device is painless.

Another barrier to PO use was the increased workload, particularly reported in Burkina Faso. This burden was especially evident in high-volume settings with limited staff, making PO use challenging due to time constraints, particularly during peak periods or when dealing with impatient patients. To alleviate this burden, some PHCs implemented strategies such as separating adult and child consultations, using the PO during triage, or conducting paediatric consultations with two HCWs. However, the feasibility of these solutions remained dependent on sufficient human resources.

### PO adoption

Two configurations were identified as pathways to adoption. The first configuration centred on enhanced diagnostic confidence among HCWs. Prior to the introduction of the PO, many described challenges in making accurate diagnoses: “Before, we consulted children with uncertainty” (Mali, T1, PHC1). In the absence of adequate equipment, diagnoses often relied on individual judgment, leading to difficult decisions – for example, whether to keep a sick child for observation or send them home despite signs of worsening health. Introducing the PO reduced this uncertainty, providing HCWs with a greater sense of confidence and calm in their decision-making: “The PO allows me to be sure of what I’m doing and saves lives” (Mali, T2, PHC4).

The second configuration was linked to HCWs’ direct observation of the PO’s usefulness in improving childcare, which gave them a sense of relief and motivation to adopt the device in their daily practice. This response was particularly noted in Mali. Although the benefits of the PO were often inferred due to the limited availability of oxygen at the decentralized level, HCWs reported feeling reassured when they received updates about a child’s improved condition, either through caregivers’ feedback or follow-up exchanges with hospital staff.

### Structural and institutional barriers to PO adoption

While many positive configurations supporting the use and adoption of the PO were identified, our interviews, particularly with institutional-level respondents, highlighted several structural and institutional factors that could undermine these dynamics and hinder long-term adoption.

Adoption often depends on HCWs’ ability to observe the benefits of the PO in practice. However, structural limitations related to follow-up care and oxygen delivery services can restrict this experience: “If you don’t have the ability to provide ventilation for the child on-site… the problem is not solved.” (Guinea, institutional actor). Since oxygen therapy is typically only available in hospital settings, HCWs must rely on patient transfers, which can be complex and poorly coordinated, especially following the end of the AIRE project. One HCW explained: “it’s not the same as when the investigators were around.” (Niger, T2, PHC1). In the interviews, the end of project supervision—previously valued for its monitoring and reminders— was often perceived as a threat to the sustained use of the PO, as ongoing support mechanisms will no longer be available.

Potential barriers also include the relatively low prevalence of hypoxemia within the total volume of consultations observed in decentralized settings compared to hospital environments. Some HCWs reported rarely encountering hypoxemia cases, which may reduce confidence in the diagnostic value of the PO over time.

Additionally, while this study focused on individual adoption, interviews revealed that sustainability also depends on institutional integration, which remains limited due to resource constraints. Despite signs of individual adoption, PHCs often showed passivity toward institutionalizing the PO, relying instead on NGOs to sustain its integration through continued project support or new initiatives.

Institutional challenges were further exacerbated by staff turnover and insufficient training for newly recruited HCWs, raising concerns about the continuity of PO use. As one institutional actor warned, “In three years, none [of the staff] will be trained to use the PO” (Burkina Faso, institutional actor). Similarly, the absence of maintenance and replacement systems for broken devices poses another barrier to sustainability, as illustrated by a respondent in Niger who stated, “the care of the child with the pulse oximeter will stop… and we have no way to bring in a new device” (Niger, institutional actor).

## Discussion

This study highlights that individual adoption of the PO in PHCs was relatively high, a trend further supported by quantitative data from the AIRE project, which confirmed high utilization rates [38]. However, the intervention produced four distinct outcomes regarding the use and adoption of the PO. Non-utilization was primarily observed in high-workload settings and in cases where the device could not be used on agitated children. Mechanical use occurred when external pressures, such as supervision, motivated HCWs to use the device out of obligation or constraint, raising concerns about sustainability. Utilization was linked with increased awareness of the PO’s usefulness, supportive environments, and feelings of pride and privilege in accessing the device. Finally, adoption reflected strengthened diagnostic confidence and a sense of pride and relief from contributing to improved care for children.

Several contextual elements shaped responses to the intervention and influenced PO use. Being trained emerged as central, not only for mastering the tool but, more importantly, for recognizing its practical utility. Without multiple waves of training and formative supervision, the use of PO could have been negatively affected. As shown in other studies, limited training risks undermining collective ownership and shared responsibility [39]. Quantitative data from another AIRE study confirmed that training is strongly associated with PO acceptability [38].

The decentralized implementation of the intervention also influenced HCWs’ perceptions. In settings with limited diagnostic tools, the PO was often regarded as a valuable innovation, enhancing confidence in diagnosis and decision-making. This aligns with findings from a realist study, which highlighted that the scarcity of diagnostic tools can encourage the adoption of new technologies [39]. Indeed, perceptions of its value varied across settings. In Nigeria, some HCWs questioned whether the PO provided added diagnostic value or simply delayed referrals they already deemed necessary [22]. In contrast, studies conducted in settings with limited access to laboratory services and located farther from hospitals emphasized the PO’s role in assessment, stabilization, and referrals [40,41].

Supervision and external motivation also influenced responses, though the impact varied depending on how supervision was perceived. A realist evaluation in Guatemala distinguished between control-focused supervision, which emphasized pressure to meet targets, and support-focused supervision, which prioritized encouragement and practical guidance [42]. Studies have shown that leadership styles and supervisory approaches can significantly influence adoption rates. For example, in Kenya, differences in PO use were partially attributed to variations in leadership styles among senior doctors [43]. In Nigeria, the absence of external support negatively affected PO adoption rates [22]. The end of AIRE project supervision was often seen as a risk to sustained use, given the lack of established structures for ongoing support.

Practical and structural constraints also shaped responses to the intervention. Children’s agitation during measurements was widely reported as a barrier, consistent with findings from other studies [40,44]. High workloads and limited human resources complicated consistent use, reflecting challenges noted in studies on other diagnostic technologies [45,46]. In Guinea, where the number of consultations was lower during the AIRE project, these challenges were less reported [47].

The intervention elicited a range of responses among HCWs, influenced by perceptions of the PO as a valuable innovation and by different types of motivation. We have seen that the PO was widely regarded as an important tool in decentralized settings, where diagnostic technologies are often scarce. This perception motivated HCWs, who viewed access to advanced equipment as a privilege and a source of legitimacy. Previous studies have highlighted how the social positioning and roles of HCWs influence their relationship with technology [48,49]. Abejirinde et al. [39] also found that a sense of pride in being selected for a project can facilitate the adoption of innovations. This dynamic was further reinforced by increased patient trust [38], echoing findings from other studies where families’ confidence in HCWs improved after the introduction of the PO [40,43,50].

HCWs’ motivations also varied depending on whether they were externally driven or internally rooted. Insights from Self-Determination Theory [51] suggest that autonomous motivation, grounded in personal values, is generally more sustainable than controlled motivation, which depends on external pressures. In this context, HCWs’ ownership of the PO was strengthened when key actors – such as supervisors or managers – were perceived as supportive rather than controlling. Supportive interactions encouraged HCWs to see the PO as a tool to improve care, while supervision perceived as imposing constraints sometimes resulted in mechanical use without genuine engagement. Interestingly, mechanical use, initially driven by external reminders, did not necessarily hinder adoption. Over time, reliance on external motivation sometimes transitioned into voluntary adoption, reinforced by repeated positive experiences with the PO. Similar patterns were observed in Nigeria, where constrained use eventually evolved into internalized adoption as HCWs gained familiarity and confidence in the tool [33].

The PO also enhanced HCWs’ diagnostic confidence, which proved to be a key mechanism promoting adoption. HCWs reported greater certainty in their assessments, leading to faster decision-making. Comparable findings were observed in India [52], Malawi, and Bangladesh [40]. However, this effect was less pronounced in hospital settings, where HCWs already had access to more diagnostic equipment and training. Within the AIRE project, referral rates for hypoxemic cases were higher than for other severe cases without hypoxemia [53], suggesting a greater reliance on PO-based diagnostics. Similar trends were noted in Malawi [41] and India [52].

Beyond diagnostic confidence, HCWs described feelings of pride and relief when observing the PO’s positive effects on patient care, further motivating its adoption. Seeing tangible benefits reinforced their belief in the device’s utility, aligning with the concepts of observability [34] and result demonstrability [54]. These concepts emphasize that the ability to observe and measure a tool’s impact encourages its adoption. A realist review similarly identified the ability to observe positive outcomes as a key motivator for HCWs to use oxygen [55].

Finally, our findings also highlight the importance of addressing structural and institutional challenges to ensure the long-term adoption and sustainability of the PO. While improved diagnostics enhanced decision-making, limited access to treatment and the need for emergency hospital referrals for oxygen therapy could demotivate HCWs, diminishing the perceived effectiveness of the PO. Well-documented barriers to hospital referrals—including distance, poor road conditions, lack of transportation, and family costs—further restricted access to care [56,57]. The highest number of referrals during the project was reported in Niger, likely due to the close proximity of PHCs to hospitals, particularly in the capital [58]. Additional challenges included supply chain issues, device maintenance, and training gaps for newly recruited staff. Concerns about the absence of maintenance systems and replacement devices raised fears of disruptions in patient care, echoing findings from other studies that identified malfunctioning devices and insufficient training as major obstacles to technology adoption [40,43,59]. Despite encouraging signs of individual adoption, ensuring the sustainability of the PO requires institutional commitment to overcome these barriers. This includes formalizing its use in clinical protocols, establishing regular training programs to address staff turnover, developing reliable supply chains for maintenance and replacement and support hospital referrals and oxygen therapy delivery. Such measures are critical to prevent these challenges from undermining adoption efforts and to support the integration of the PO into health systems effectively.

This study has some limitations that should be acknowledged. First, contextual and methodological biases may have influenced the findings. Social desirability bias [60] could have led HCWs to present an idealized image of their work, particularly given their dependency relationships with NGOs and donors [61]. Similar pressures to demonstrate success and secure funding are well-documented in the global health sector [62]. Additionally, practical and time constraints posed challenges to conducting realist interviews [63], partly due to limited training time for investigators and perceived power dynamics between interviewers and participants. Such challenges have been highlighted in other studies [64]. To mitigate these limitations, several strategies were implemented. Investigators explicitly clarified their independence from the NGOs involved, encouraging participants to speak freely and anonymously. Furthermore, triangulation of data sources was used to cross-verify information and enhance the reliability of the results.

Despite these limitations, this study provides one of the first applications of realist evaluation to explore the individual adoption of a technological innovation in primary health care settings. It offers a solid foundation for future research on the dynamics of technology adoption in similar contexts.

## Conclusion

This study suggests that POs are generally used and even adopted by HCWs in decentralized contexts, especially where diagnostic tools are scarce, and their benefits for patient care are observable. Its use enhances diagnostic confidence and reinforces perceptions of its utility. However, structural and institutional challenges, including supply, maintenance, staff training, and access to hospital for oxygen treatment, may threaten its long-term integration into health systems. Addressing these barriers will be essential to ensure the sustainability of the PO and its impact on child health outcomes.

## ACKNOWLEDGMENTS

We thank all the participants of this study, as well as the healthcare staff involved in the AIRE project. We also express our gratitude to the field project staff and the AIRE Research Study Group. Finally, we thank the Ministries of Health of the participating countries for their support.

## The AIRE Research Study Group: Country investigators

Ouagadougou, Burkina Faso: S. Yugbaré Ouédraogo (PI), V. M. Sanon Zombré (CoPI), Conakry, Guinea: M. Sama Cherif (CoPI), I. S. Diallo (CoPI), D. F. Kaba, (PI). Bamako, Mali: A. A. Diakité (PI), A. Sidibé, (CoPI). Niamey, Niger: H. Abarry Souleymane (CoPI), F. Tidjani Issagana Dikouma (PI). **Research coordinators & data centers: Inserm U1295, Toulouse 3 University, France:** H. Agbeci (Int Health Economist), L. Catala (Research associate), D. L. Dahourou (Research associate), S. Desmonde (Research associate), E. Gres (PhD Student), G. B. Hedible (Int research project manager), V. Leroy (research coordinator), L. Peters Bokol (Int clinical research monitor), J. Tavarez (Research project assistant), Z. Zair (Statistician, Data scientist). **CEPED, IRD, Paris, France:** S. Louart (process manager), V. Ridde (process coordination). **Inserm U1137, Paris, France:** A. Cousien (Research associate). **Inserm U1219**, EMR271 IRD, **Bordeaux University, France**: R. Becquet (Research associate), V. Briand (Research associate), V. Journot (Research associate). **PACCI, CHU Treichville, Abidjan, Côte d’Ivoire**: S. Lenaud (Int data manager), C. N’Chot (Research associate), B. Seri (Supervisor IT), C. Yao (data manager supervisor). **Consortium NGOs partners: Alima-HQ (consortium lead), Dakar, Sénégal**: G. Anago (Int Monitoring Evaluation Accountability And Learning Officer), D. Badiane (Supply chain manager), M. Kinda (Director), D. Neboua (Medical officer), P. S. Dia (Supply chain manager), S. Shepherd (referent NGO), N. di Mauro (Operations support officer), G. Noël (Knowledge broker), K. Nyoka (Communication and advocacy officer), W. Taokreo (Finance manager), O. B. Coulidiati Lompo (Finance manager), M. Vignon (Project Manager). **Alima, Conakry, Guinea:** P. Aba (clinical supervisor), N. Diallo (clinical supervisor), M. Ngaradoum (Medical Team Leader), S. Léno (data collector), A. T. Sow (data collector), A. Baldé (data collector), A. Soumah (data collector), B. Baldé (data collector), F. Bah (data collector), K. C. Millimouno (data collector), M. Haba (data collector), M. Bah (data collector), M. Soumah (data collector), M. Guilavogui (data collector), M. N. Sylla (data collector), S. Diallo (data collector), S. F. Dounfangadouno (data collector), T. I. Bah (data collector), S. Sani (data collector), C. Gnongoue (Monitoring Evaluation Accountability And Learning Officer), S. Gaye (Monitoring Evaluation Accountability And Learning Officer), J. P. Y. Guilavogui (Clinical Research Assistant), A. O. Touré (Country health economist), J. S. Kolié (Country clinical research monitor), A. S. Savadogo (country project manager). **Alima, Bamako, Mali:** F. Sangala (Medical Team Leader), M. Traore (Clinical supervisor), T. Konare (Clinical supervisor), A. Coulibaly (Country health economist), A. Keita (data collector), D. Diarra (data collector), H. Traoré (data collector), I. Sangaré (data collector), I. Koné (data collector), M. Traoré (data collector), S. Diarra (data collector), V. Opoue (Monitoring Evaluation Accountability And Learning Officer), F. K. Keita (medical coordinator), M. Dougabka (Clinical research assistant then Monitoring Evaluation Accountability And Learning Officer), B. Dembélé (data collector then Clinical research assistant), M. S. Doumbia (country health economist), G. D. Kargougou (country clinical research monitor), S. Keita (country project manager). **Solthis-HQ, Paris**: S. Bouille (NGO referent), S. Calmettes (NGO referent), F. Lamontagne (NGO referent). **Solthis,** Niamey: K. H. Harouna (clinical supervisor), B. Moutari (clinical supervisor), I. Issaka (clinical supervisor), S. O. Assoumane (clinical supervisor), S. Dioiri (Medical Team Leader), M. Sidi (data collector), K. Sani Alio (Country supply chain officer), S. Amina (data collector), R. Agbokou (Clinical research assistant), M. G. Hamidou (Clinical Research Assistant), S. M. Sani (Country health economist), A. Mahamane, Aboubacar Abdou (data collector), B. Ousmane (data collector), I Kabirou (data collector), I. Mahaman (data collector), I Mamoudou (data collector), M. Baguido (data collector), R. Abdoul (data collector), A. Sahabi (data collector), F. Seini (data collector), Z. Hamani (data collector), L-Y B Meda (Country clinical research monitor), Mactar Niome (country project manager), X. Toviho (Monitoring Evaluation Accountability And Learning Officer), I. Sanouna (Monitoring Evaluation Accountability And Learning Officer), P. Kouam (program officer). **Terre des hommes-HQ, Lausanne:** S. Busière (NGO referent), F. Triclin (NGO referent). **Terre des hommes, BF:** A. Hema (country project manager), M. Bayala (IeDA IT), L. Tapsoba (Monitoring Evaluation Accountability And Learning Officer), J. B. Yaro (Clinical research assistant), S. Sougue (Clinical research assistant), R. Bakyono (Country health economist), A. G. Sawadogo (Country clinical research monitor), A. Soumah (data collector), Y. A. Lompo (data collector), B. Malgoubri (data collector), F. Douamba (data collector), G. Sore (data collector), L. Wangraoua (data collector), S. Yamponi (data collector), S. I. Bayala (data collector), S. Tiegna (data collector), S. Kam (data collector), S. Yoda (data collector), M. Karantao (data collector), D. F. Barry (Clinical supervisor), O. Sanou (clinical supervisor), N. Nacoulma (Medical Team Leader), N. Semde (clinical supervisor), I. Ouattara (Clinical supervisor), F. Wango (clinical supervisor), Z. Gneissien (clinical supervisor), H. Congo (clinical supervisor). **Terre des hommes, Mali:** Y. Diarra (clinical supervisor), B. Ouattara (clinical supervisor), A. Maiga (data collector), F. Diabate (data collector), O. Goita (data collector), S. Gana (data collector), S. Diallo (data collector), S. Sylla (data collector), D. Coulibaly (Tdh project manager), N. Sakho (NGO referent). **Country SHS team: Burkina Faso**: K. Kadio (consultant and research associate), J. Yougbaré (data collector), D. Zongo (data collector), S. Tougouma (data collector), A. Dicko (data collector), Z. Nanema (data collector), I. Balima (data collector), A. Ouedraogo (data collector), A. Ouattara (data collector), S. E. Coulibaly (data collector). **Guinea**: H. Baldé (consultant and research associate), L. Barry (data collector), E. Duparc Haba (data collector). **Mali**: A. Coulibaly (consultant and research associate), T. Sidibe (data collector), Y. Sangare (data collector), B. Traore (data collector), Y. Diarra (data collector). **Niger**: A. E. Dagobi (consultant and research associate), S. Salifou (data collector), B. Gana Moustapha Chétima (data collector), I. H. Abdou (data collector)

## FOOTNOTES

### Contributors

All authors contributed to the conceptualization of the research. HB, AC, AED, and KK conducted data collection with support from SL and the AIRE research study group. SL, HB, AC, AED, and KK carried out the data analysis. SL prepared the initial draft of this article. All authors participated in data interpretation and reviewed the final manuscript.

### Ethics approval and consent to participate

Ethics approval and consent to participate The AIRE research protocol, the information notice (translated in vernacular languages), the written consent form and any other relevant document have been submitted to each national ethics committee, to the Inserm Institutional Evaluation Ethics Committee (IEEC) and to the WHO Ethics Review Committee (WHOIZIERC). All the aforementioned ethical committees reviewed and approved the protocol and other key documents (Comité d’Ethique pour la Recherche en Santé (CERS), Burkina Faso n°2020–4IZI070; Comité National d’Ethique pour la Recherche en Santé (CNERS), Guinea n°169/CNERS/21; Comité National d’Éthique pour la Santé et les Sciences de la vie (CNESS), Mali n°127/MSDSIZICNESS; Comité National d’Ethique pour la Recherche en Santé (CNERS) Niger n°67/2020/CNERS; Inserm IEEC n°20–720; WHOIZIERC n° ERC.0003364). This study has been retrospectively registered by the Pan African Clinical Trials Registry on June 15^th^, 2022, under the following Trial registration number: PACTR202206525204526.

### Data Availability Statement

The datasets generated and analysed during the current study are not publicly available. Access to processed deidentified participant data will be made available to any third Party after the publication of the main AIRE results stated in the Pan African Clinical Trial Registry Study statement (PACTR202206525204526, registered on 06/15/2022), upon a motivated request (concept sheet), and after the written consent of the AIRE research coordinator (Valériane Leroy, Inserm U1295 Toulouse, France, orcid.org/0000-0003-3542-8616) obtained after the approval of the AIRE publication committee, if still active.

