## Appendix for "Understanding health innovation adoption: A realist evaluation of pulse oximeter implementation in primary care for children under five in four West African countries"

1 – First modelling of the program theory intervention

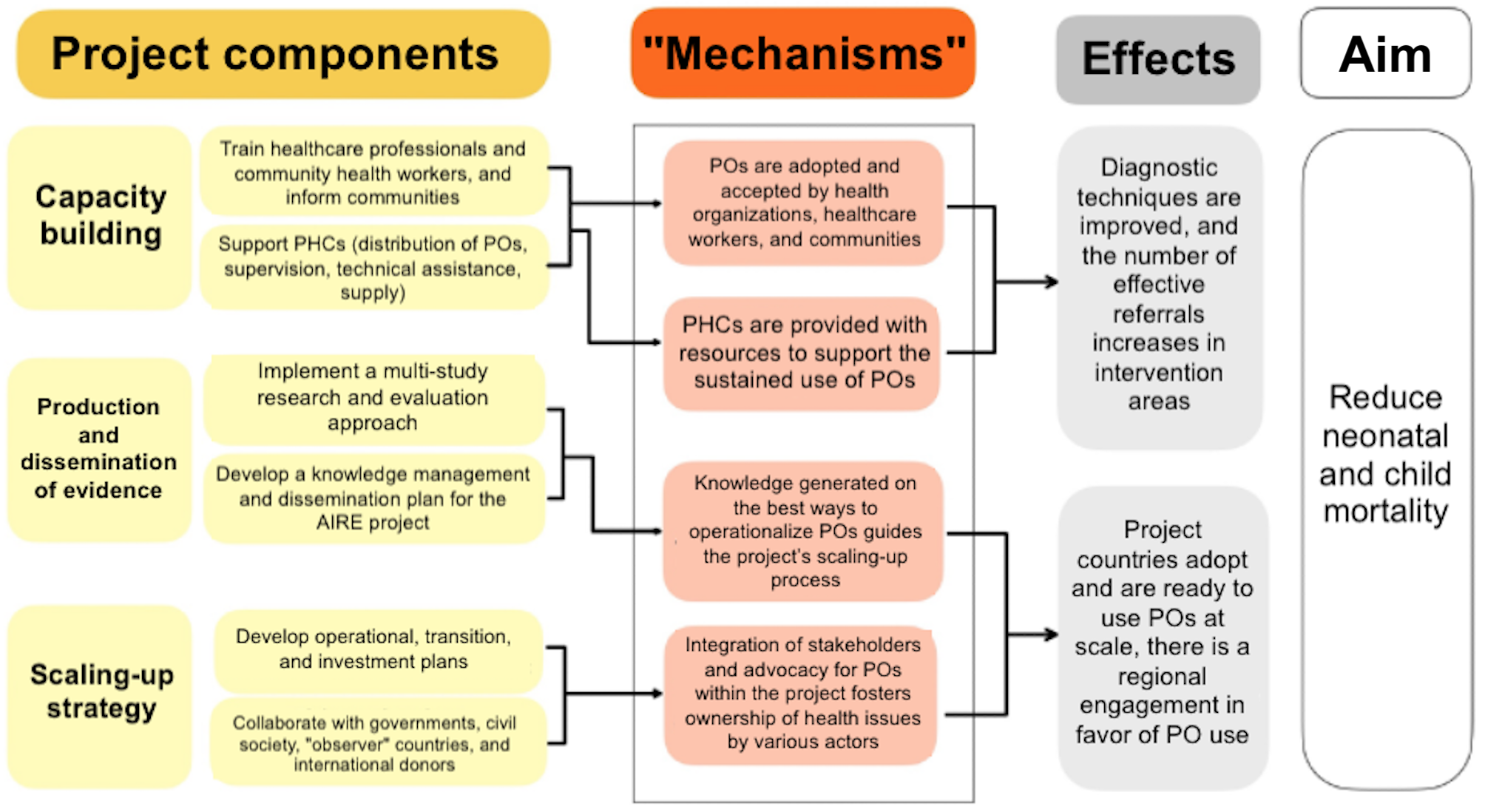

2 – Refined intervention theory

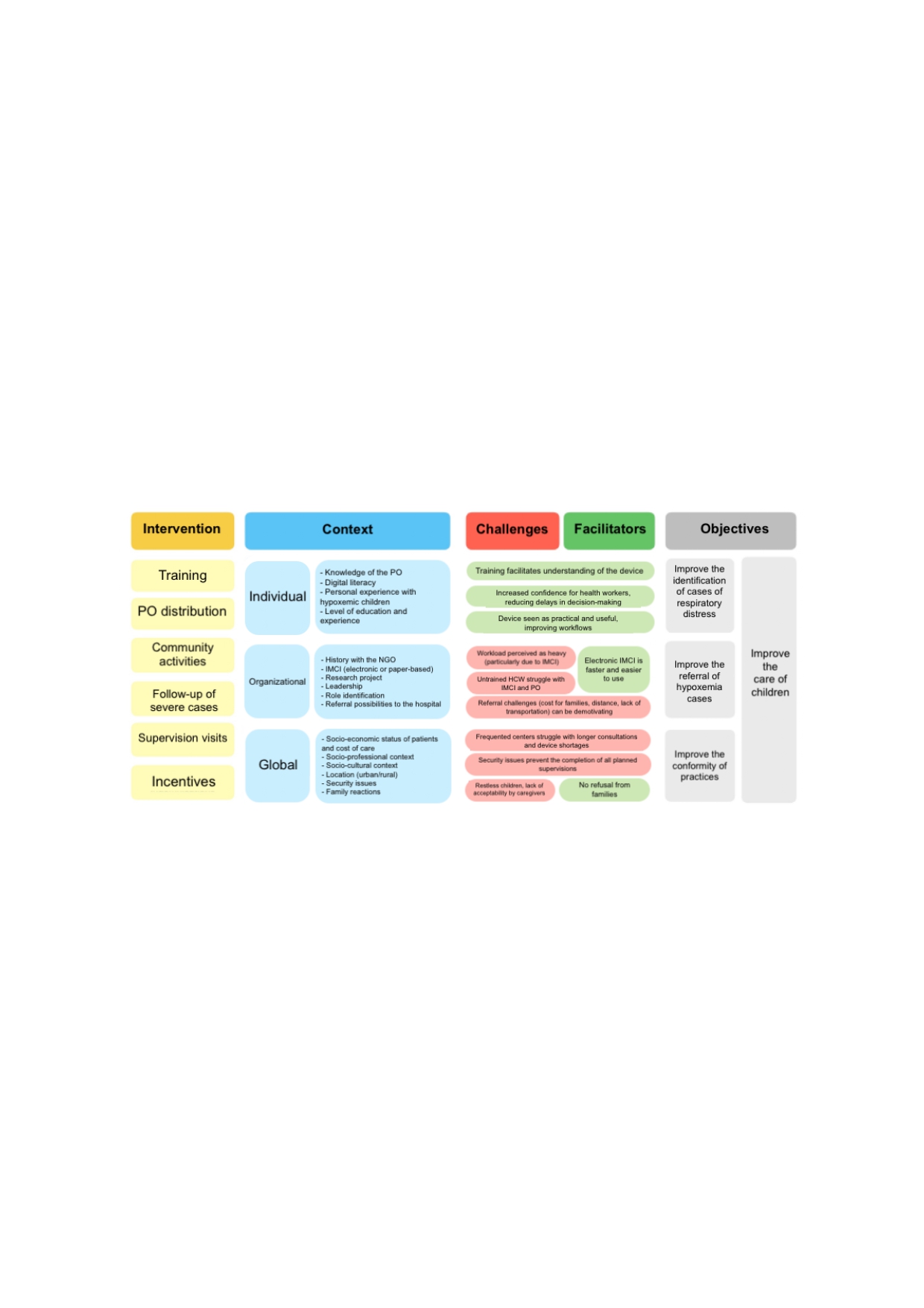

3 A – Final list of hypotheses regarding PO use
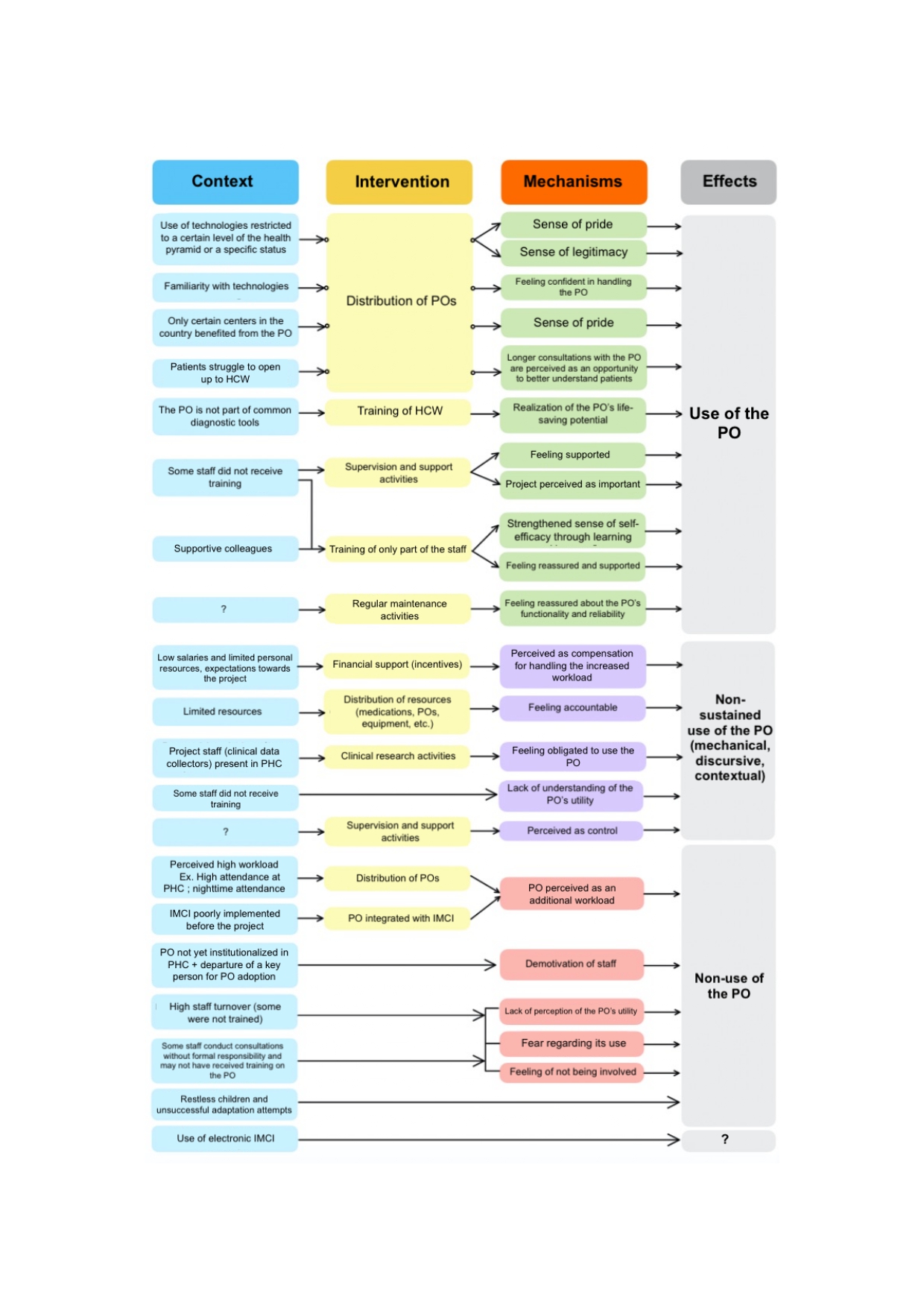

3 B - Final list of hypotheses regarding PO adoption

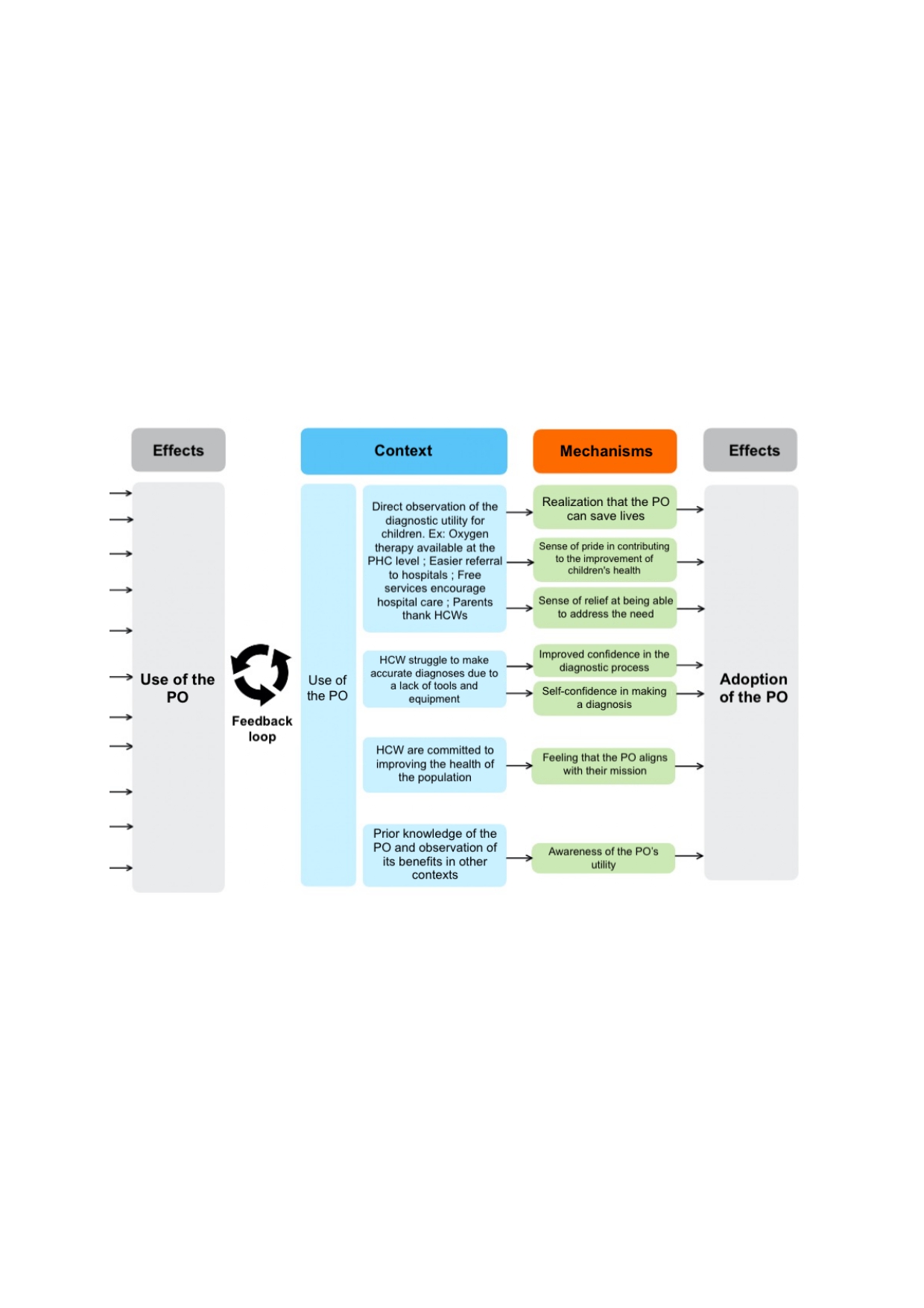
